# A study protocol for a randomised controlled trial evaluating the safety and efficiency of the YEARS algorithm versus computed tomography pulmonary angiography only for suspected acute pulmonary embolism in patients with cancer: the Hydra Study

**DOI:** 10.64898/2026.01.14.26344055

**Authors:** E.S.L. Martens, B. Akerboom, C. Baumgartner, R.E. Brouwer, C. Cavallaro, M. Coppens, G. Costantino, F. Couturaud, A. D’Errico, Y.P.A. Dooren, F. Gianni, R. van der Griend, M.J.J.H. Grootenboers, O. Hugli, T. van der Hulle, Z. Jaderi, D. Jimenez, P.W. Kamphuisen, V.R. Lanting, J. Leentjens, M.L. Maas, I. Mahé, O.A. van Meer, T.E. van Mens, M. Out, R. Pola, D. Pulver, J. Raskin, M. Righini, R.A. Sprenger, M.A.M. Stals, R. Talerico, T. Tritschler, M. ten Wolde, F.A. Klok, M.V. Huisman, the HYDRA study group

## Abstract

**Background:** Pulmonary embolism (PE) is frequently suspected in patients with cancer due to non-specific symptoms suggestive of PE and their inherent risk of venous thromboembolism (VTE). Commonly used clinical decision rules (CDRs) and D-dimer testing may be less reliable in this specific population. Due to the lack of guideline recommendations on the optimal diagnostic approach and the perceived futility of D-dimer in patients with cancer, clinicians often use computed tomography pulmonary angiography (CTPA) as a sole diagnostic test. This practice exposes patients to potentially unnecessary radiation and harm, and contributes to a significant burden on healthcare systems through inefficient resource allocation. The YEARS algorithm is an easy-to-use CDR that has been shown to safely exclude PE without the need for CTPA in a diverse population of patients with suspected PE.

**Objective:** To compare the safety and efficiency of the YEARS algorithm to the safety and efficiency of CTPA only in the diagnostic management of acute PE in cancer patients.

**Design and interventions:** The Hydra study (ClinicalTrials.gov NCT04657120) is an investigator-initiated, multicentre, multinational open-label, randomised, non-inferiority trial with blinded adjudication of outcome events, comparing the YEARS algorithm with CTPA only in the diagnostic work-up of cancer patients with clinically suspected acute PE. Participants are randomised in a 1:1 ratio to each diagnostic strategy via a web-based system. The trial anticipates to include 1566 patients.

**Participants:** Consecutive patients with active cancer who are hospitalized or present to the emergency department, outpatient clinic, or thrombosis clinic with clinically suspected PE and who are not receiving therapeutic anticoagulation or have an indication for anticoagulation therapy other than PE.

**Study outcomes:** The primary safety outcome is the proportion of symptomatic and objectively proven fatal or non-fatal VTE (i.e., PE or deep vein thrombosis in the upper or lower extremities) or death with undetermined cause where acute PE could not be ruled out as contributing factor during three months follow-up in patients in whom PE was ruled out at initial testing. The primary efficiency outcome is the proportion of negative CTPA scans for PE, relative to the total number of CTPA scans performed at initial testing.

**Implication:** This trial will provide pivotal data on the optimal diagnostic approach for suspected acute PE in patients with cancer.

**Strengths and limitations of this study:**

- The Hydra study is the first prospective, randomised trial comparing the safety and efficiency of the YEARS algorithm with CTPA only for suspected acute PE in patients with active cancer.
- The enrolment of a large number of patients from multiple teaching and general hospitals across various clinical settings and countries enhances the generalisability of this study.
- The randomised design and the vulnerable study population may render the study challenging.

## Background and rationale

Pulmonary embolism (PE), together with deep vein thrombosis (DVT) referred to as venous thromboembolism (VTE), is a well-known and serious complication in patients with cancer.^1^ The diagnostic standard imaging tool for diagnosing PE is computed tomography pulmonary angiography (CTPA), valued for its high accuracy, short acquisition time, and wide availability.^2^ However, concerns about unnecessary imaging and the potential harms associated with the overuse of CTPA, such as radiation exposure and incidental non-PE findings of unclear significance, have led to the development of clinical decision rules (CDRs), such as the YEARS algorithm^3^, as complementary diagnostic tests.^4–6^

While the utility of CDRs combined with D-dimer testing is well established in the diagnostic work-up of non-cancer patients suspected of PE, their safety and efficiency in patients with cancer remains debated.^7^ This is primarily due to the lower specificity of D-dimer in the presence of cancer, as cancer itself may lead to elevated D-dimer levels in absence of thrombosis. Previous studies have shown that the incidence of normal D-dimer levels (cut off at 500 ng per millilitre or age-adjusted) in patients with cancer and suspected PE may be as low as 10% to 15%.^8,9^ A meta-analysis of individual patient data from 16 studies, including 2,219 patients with active cancer and suspected PE (11%), showed that imaging could be avoided in 21% of patients following application of the YEARS algorithm. In comparison, the predicted efficiency for other CDRs using fixed and adjusted D-dimer thresholds ranged from 9.6% to 26%.^6^ In the prospective YEARS study, the safety and efficiency of the YEARS algorithm in consecutive in- and outpatients with clinically suspected acute PE was studied. PE was excluded without CTPA in 18% of the subgroup of 336 patients with underlying cancer on the basis of a negative YEARS algorithm. At three months, 2.6% of these patients developed VTE, compared to 0.8% in the total cohort.^3^ Nonetheless, given the inherently high thromboembolic risk in patients with cancer, it is possible that some of the VTE events at follow-up were new events rather than missed diagnoses. The persistent uncertainty regarding the accuracy and diagnostic performance of CDR and D-dimer testing in patients with cancer likely contributes to the wide variation of diagnostic management in clinical practice. In the absence of well-designed diagnostic studies focusing on patients with cancer and suspected PE, clinicians often opt for CTPA as the primary diagnostic tool out of concern that they might miss a diagnosis of PE and the presumed futility of D-dimer as a diagnostic test. Given the increased risk of bleeding in patients with cancer, especially during the course of anticoagulation therapy, a diagnostic algorithm that can safely rule out PE without the need for CTPA in these patients will improve patient care by reducing the detection and subsequent unnecessary overtreatment of false-positive radiologic findings of (subsegmental) PE and incidental findings of unclear significance.^10–12^ Moreover, it will also considerably lower the burden on healthcare systems and costs due to inefficient resource allocation.^3,13^ We hypothesize that the YEARS algorithm is non-inferior to CTPA only with regard to three-month VTE rates (safety indicator) and will reduce the number of unnecessary CTPA scans and anticoagulation therapy in patients with clinically suspected PE and active cancer (efficiency indicator).

### Study overview

#### Study design and objectives

The Hydra study (ClinicalTrials.gov Identifier: NCT04657120) is an investigator-initiated, multicentre, multinational open-label, randomised, non-inferiority, trial with blinded adjudication of outcome events. Patients will be recruited from teaching and general hospitals across the Netherlands, Belgium, France, Italy, Spain and Switzerland.

The primary objective of the Hydra study is to assess whether diagnostic management according to the YEARS algorithm is non-inferior in terms of safety to CTPA only in ruling out PE in patients with active cancer suspected of acute PE. A secondary objective is to assess whether the YEARS algorithm is superior in terms of efficiency, if non-inferiority is confirmed. Additional objectives are to describe differences in i) the detection of isolated subsegmental PE at initial testing, ii) the timing of symptomatic VTE during follow-up iii) the occurrence of incident VTE during follow-up, iv) the occurrence of contrast-induced complications (e.g., allergic reactions, contrast-induced nephropathy), v) practice patterns of anticoagulation therapy and their safety in terms of recurrent VTE and major bleeding, vi) quality of life in patients with PE at initial testing and follow-up, as measured by the Pulmonary Embolism Quality of Life (PEmb-QoL) questionnaire^14^, vii) the performance of the 4-Level Pulmonary Embolism Clinical Probability Score (4-PEPS)^15^ in patients randomised for the YEARS algorithm, and to describe practice patterns of anticoagulation therapy during end-of-life care in terminally ill patients.

#### Patient population and eligibility

Patients with active cancer (see **Table 1** for definitions and study eligibility criteria) who are hospitalized or present to the emergency department, outpatient clinic, or thrombosis clinic with clinically suspected PE are considered for inclusion in the trial.

**Table 1.** Study eligibility criteria.

| Inclusion criteria | Exclusion criteria |
| --- | --- |
| 1) Clinically suspected PE as judged by the attending physician<br>2) Active cancer, defined as any malignancy (excluding basal-cell and squamous-cell carcinoma of the skin), characterized by one or more of the following:<br>- diagnosed within 6 months prior to eligibility assessment (confirmed histologically or strongly suspected by the clinician)<br>- ongoing treatment for cancer or anticancer treatment within the past 6 months prior to eligibility assessment<br>- presence of metastases, including recurrent or local metastatic disease<br>3) Age $\geq 18$ years<br>4) Signed written informed consent | 1) Symptoms, suspected for PE, for $> 10$ days<br>2) Medical or psychological condition that would not permit completion of the study or signing of informed consent, including life expectancy less than 3 months, or unwillingness to sign informed consent<br>3) Treatment with full-dose therapeutic anticoagulation therapy:<br>- if initiated 24 hours or more prior to eligibility assessment (prophylactic-dose LMWH or DOAC is permitted), or;<br>- if initiation is expected for a different indication (e.g., atrial fibrillation)<br>4) Contraindication to CTPA<br>- contrast allergy<br>- impaired kidney function (eGFR $<30$ ml/min/$1.73\text{m}^2$)<br>- pregnancy<br>5) Haemodynamic instability at presentation (as a consequence of concurrent acute PE), indicated by at |
|  | least one of the following:<br>- SBP < 100 mmHg,<br>or HR >120 b/min (unless arrhythmia),<br>or SBP drop by > 40 mmHg for > 15 min.<br>- need for catecholamines to maintain adequate organ perfusion and a SBP of > 100 mmHg<br>- need for cardiopulmonary resuscitation<br>6) Suggestion of PE on previously performed CT scan, for which now PE-specific diagnostic testing is only performed as means of verification<br>7) Participating in another concurrent study on thromboprophylaxis<br>8) Prior participation in the Hydra study |

*Abbreviations: PE, pulmonary embolism; LMWH, low-molecular-weight heparin; DOAC, direct oral anticoagulant; CTPA, computed tomography pulmonary angiography; eGFR, estimated glomerular filtration rate; SBP, systolic blood pressure; mmHg, millimeters of mercury; HR, heart rate; b/min, beats per minute; CT, computed tomography.*

#### Randomisation, blinding and treatment allocation

After written informed consent is obtained, eligible patients are randomly allocated, in a 1:1 ratio, to diagnostic management by either the YEARS algorithm or CTPA only. Randomisation is performed centrally with the use of an interactive Web-response platform (Castor Electronic Data Capture) with stratification by trial centre, with concealed sequence of randomisation allocation. This is an open-label study, meaning that patients and treating physicians will not be blinded to the intervention; however, outcome assessors will be.

#### Study intervention, control-arm and follow-up

The study flow diagram is shown in **Figure 1**. If a study patient is allocated to the YEARS algorithm (i.e., experimental arm), the attending physician assesses the presence or absence of the YEARS items – being symptomatic DVT, haemoptysis, and whether PE is the most likely diagnosis – with the pretest probability dependent threshold of the D-dimer test.^3^ D-dimer concentrations are measured upon allocation to the YEARS algorithm, with automated, well-validated highly sensitive quantitative D-dimer assays according to local practice (e.g., Vidas D-dimer, STA-LIA, Innovance, Tina-Quant D-Dimer, Siemens). In patients with no YEARS items and a D-dimer concentration less than 1000 ng per millilitre (i.e., 1.0 mg per litre), PE is considered excluded and further testing is withheld. In patients with one or more YEARS items and a D-dimer concentration less than 500 ng per millilitre (i.e., 0.5 mg per litre), PE is also considered excluded and further testing is withheld. All other patients – either with no YEARS item and a D-dimer concentration of 1000 ng per millilitre (i.e., 1.0 mg per litre) or more, or with one or more items and a concentration of 500 ng per millilitre (i.e., 0.5 mg per litre) or more – are referred for CTPA to confirm or rule out the diagnosis of PE.

**Figure 1.**
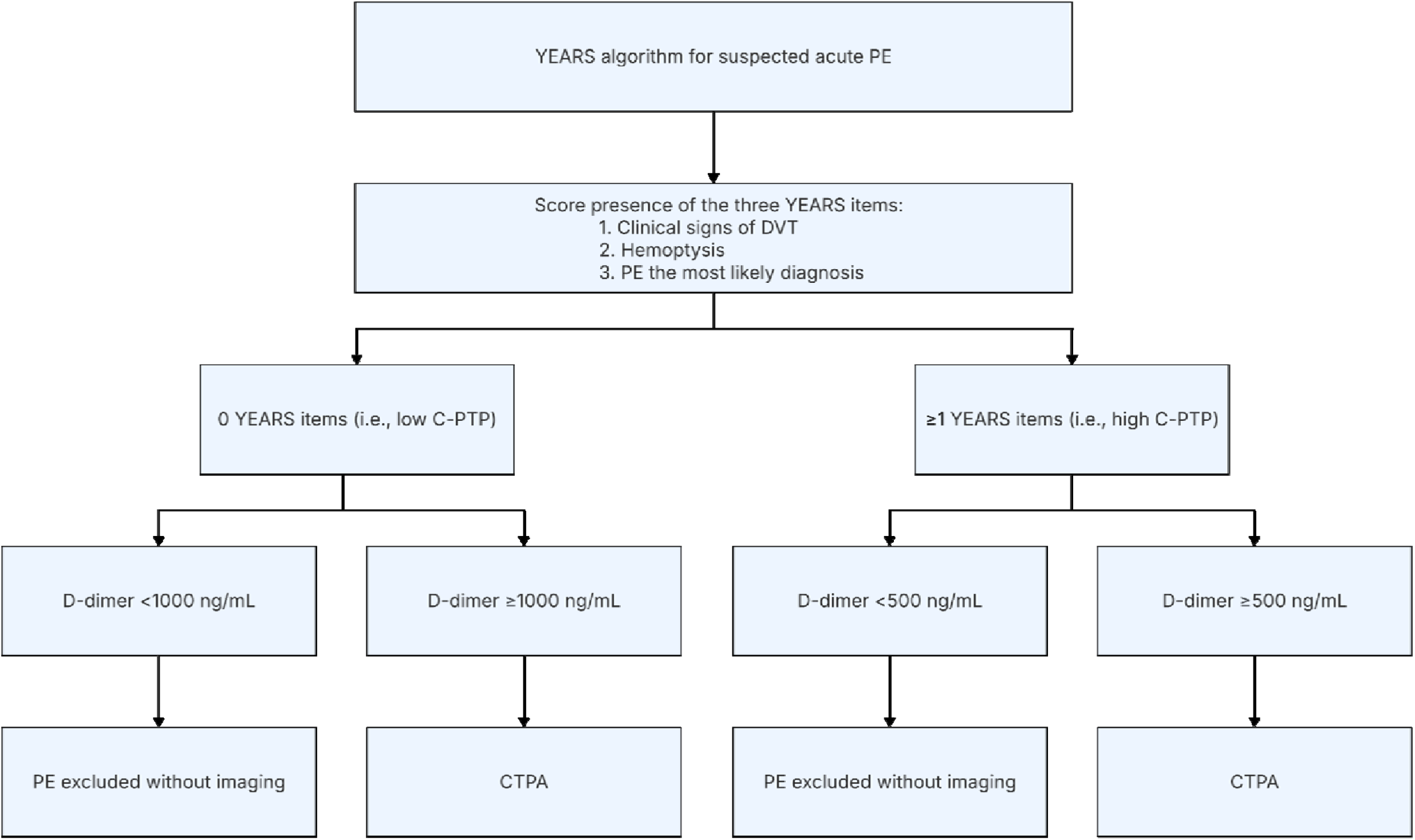
YEARS algorithm. Abbreviations: PE, pulmonary embolism; DVT, deep vein thrombosis; C-PTP, clinical pre-test probability; CTPA, computed tomography pulmonary angiography.

Study participants allocated to the control arm undergo CTPA as a standalone diagnostic test, performed according to standard local clinical practice.^16^ No routine D-dimer testing is conducted in this group.

At the time PE is diagnosed at initial testing or during the 3-month follow-up period, the decision for anticoagulation treatment (i.e., initiation, type and dosing) is left to the discretion of the treating physician. Regardless of confirmation of acute PE, all patients will receive a follow-up phone call from study personnel at 90 days (± 7 days) after the initial evaluation. Patients are instructed to contact the treating physician in case of signs or symptoms of VTE recurrence during follow-up. In addition, patients with confirmed PE at initial testing or during follow-up are requested to complete the PEmb-QoL questionnaire at the end of the follow-up period as instrument to assess quality of life following PE.^14^ In case a patient is lost to follow-up or dies during the follow-up period, study personnel will contact the family physician or clinicians involved in the patient’s care, such as those in nursing homes, hospices or other end-of-life care settings, to gather additional information.

#### Study outcomes

The primary safety outcome, also referred to as the diagnostic failure rate, is defined as the proportion of symptomatic and objectively proven fatal or non-fatal VTE (i.e., PE or DVT in the upper or lower extremities) or deaths with undetermined cause where acute PE could not be ruled out as contributing factor during three months follow-up in patients in whom PE was ruled out at initial testing (i.e., based on a negative result of the YEARS algorithm or a negative CTPA), and in whom therapeutic anticoagulation was not started (see **Supplemental Material** for the adjudication criteria).

The primary efficiency outcome is defined as the proportion of negative CTPA scans (i.e., ruling out PE), relative to the total number of CTPA scans performed during initial testing. Secondary outcomes include: (i) the occurrence of isolated subsegmental PE at baseline; (ii) incidental VTE (defined as PE or DVT that is detected by means of imaging tests performed for reasons other than clinical suspicion of VTE during follow-up) diagnosed during follow-up; (iii) major bleeding (defined according to ISTH-criteria^17,18^) occurring either with or without anticoagulation therapy; (iv) anticoagulation practice patterns during end-of-life care in terminally ill patients with cancer; (v) contrast-induced complications (i.e., contrast-induced nephropathy and allergic reactions; and (vi) quality of life following confirmed PE as measured by the PEmb-QoL questionnaire.^14^ Contrast-induced nephropathy is defined as impairment of renal function gauged as a 25% rise in serum creatinine from baseline or an increase of 0.5 milligram per decilitre (44 micromoles per litre) in absolute serum creatinine value within 48 to 72 hours following intravenous contrast administration. Allergic reactions to intravascular iodinated contrast media are defined as non-severe skin manifestations such as a pruritus, maculopapular rash or urticaria, as well as more severe anaphylactoid responses including bronchospasm, angioedema, respiratory arrest, cardiac arrest, pulmonary oedema, convulsions, and cardiogenic shock occurring between one hour to one week after contrast administration.

#### Sample size calculation and statistical analysis

The two main outcomes – safety and efficiency – will be analysed in a hierarchical manner. First, the primary safety outcome will be assessed to determine the noninferiority of management according to the YEARS algorithm compared to CTPA only. This analysis will be performed on the per-protocol population and, as a second step, the intention-to-diagnose (ITD) population. The null hypothesis (H_0_) posits that the probability of a primary safety outcome event in the control group (π_c_ – CTPA only) and in the experimental group (π_e_ – YEARS algorithm) is identical; the alternative hypothesis (H_1_) posits that the probability of an event is higher in the experimental group than in the control group. If the upper bound of the 95% CI for the difference in the event rates between the two groups exceeds the pre-specified margin for non-inferiority (expressed as an absolute risk difference at a one-sided alpha level of 5%), the null hypothesis of non-inferiority will be rejected. Only if non-inferiority is confirmed will a two-sided superiority analysis be performed on the primary efficiency outcome.

The per-protocol population will comprise all randomised patients who meet each of the following criteria: i) PE is ruled out at initial testing based on a negative result of the YEARS algorithm (i.e., managed without imaging) or a negative CTPA result, whether in the YEARS or CTPA study arm, ii) the patient remains untreated with therapeutic anticoagulation therapy, defined as no therapeutic anticoagulation in the first 7 days after initial testing, or use of therapeutic anticoagulation for a continuous duration of 14 days or less throughout the three-month follow-up period, iii) they completed the full 3-month follow-up, except in cases of death during follow-up, iv) diagnostic procedures to rule out PE were correctly applied without any major protocol deviations. Patients will be classified as having a diagnostic failure if they experience any of the following during follow-up: symptomatic, objectively proven non-fatal PE, fatal PE, DVT in the upper or lower extremities, or death with an undetermined cause (i.e., possibly PE-related). In addition, in the ITD population, a PE diagnosed at initial testing by a CTPA that was not indicated is considered a failure of the allocated diagnostic strategy. If a patient is lost to follow-up, they will be included in a secondary analysis as having a diagnostic failure.

Based on data from the original YEARS cohort, we anticipate a failure rate of 2.6% (95% CI: 1.3%-5.2%) in both study arms. A non-inferiority margin of 2.0% was set. Assuming no difference between standard management by CTPA and management according to the YEARS algorithm, a total of 1566 patients in the per-protocol population will provide 80% power to demonstrate that the upper bound of the one-sided 95% Cl excludes a difference greater than 2.0% in favour of the CTPA group.

Patient characteristics will be summarized using their mean and standard deviation, median and interquartile range, or percentage with 95% CI, as appropriate. For the main and secondary outcome analyses, the absolute differences between the two diagnostic strategies are calculated with exact 95% CI around the observed incidences and proportions.

In response to an observed higher than anticipated all-cause mortality rate in the total study population, we decided to conduct an interim analysis once 700 patients have been randomised and have completed the 3 months follow-up. The purpose of this interim analysis is to account for the higher than anticipated mortality rate, which could be linked to a higher prevalence of PE-related death or death with an underdetermined cause, and in turn, a relevant higher incidence of the primary outcome than was used in the power analysis. Accordingly, the sample size will be recalculated based on prespecified absolute margins for defining non-inferiority (**Table 2). The inte**rim analysis will be performed by an independent statistician. Based on the observed diagnostic failure rate, adjusted alpha levels for the interim and final analysis will be determined using O’Brien-Fleming alpha spending function. If the early-trial results allow to demonstrate non-inferiority, subject recruitment will be ended.

**Table 2.** Prespecified margins for defining non-inferiority.

| **Actual observed diagnostic failure rate in the control-arm (CTPA only)** | **Margin for defining non-inferiority** |
| --- | --- |
| 2.6% – 3.5% | 2.0% |
| 3.6% – 4.5% | 2.3% |
| 4.6% – 5.5% | 2.6% |
| 5.6% – 6.5% | 2.9% |
| 6.6% – 7.5% | 3.2% |
| 7.6% – 8.5% | 3.5% |
| 8.6% – 9.5% | 3.8% |
| 9.6% – 10.5% | 4.1% |
| 10.6% – 11.5% | 4.4% |
| 11.6% – 12.5% | 4.7% |
| 12.6% – 13.5% | 5.0% |
| 13.6% – 14.5% | 5.0% |
| 14.6% – 15.5% | 5.0% |
| 15.6% – 16.5% | 5.0% |

*Abbreviation: CTPA, computed tomography pulmonary angiography.*

#### Implications and expected impact of the Hydra study

This trial will provide important data on the optimal diagnostic approach for suspected acute PE in the cancer population. With its rigorous design, the Hydra study will provide a comprehensive insight into the clinical utility of the YEARS algorithm versus CTPA as a standalone diagnostic test. More specifically:

1. The Hydra study is the first prospective, randomised trial directly comparing the safety and efficiency of the YEARS algorithm with CTPA only for suspected acute PE in patients with active cancer. This will help determine the most appropriate approach in this vulnerable population balancing the safety and efficiency of these diagnostic strategies, ultimately establishing a benchmark for the clinical management of suspected acute PE in patients with cancer.
2. The prospective design and direct comparison of both diagnostic strategies will help to improve differentiating between missed PE diagnoses and new-onset VTE, thereby improving our understanding of diagnostic performance in this high-risk population. To date, no study has specifically evaluated the diagnostic performance of CTPA as a standalone test in patients with cancer and suspected PE.
3. Finally, the Hydra study examines anticoagulation treatment decisions, associated outcomes, and disease-related quality of life following PE, thereby extending its scope beyond the diagnostic accuracy of the tested strategies. This comprehensive approach allows the trial to indirectly assess the safety of treatment decisions based on diagnostic findings and offers meaningful insight into contemporary clinical practice and its broader impact.

### Study organisation

#### Ethical considerations and regulations

The trial is being conducted in accordance with globally accepted standards of the International Council for Harmonization on Good Clinical Practice, in alignment with the latest revision of the ethical principles outlined in the Declaration of Helsinki and in compliance with applicable local regulations.

The protocol, any substantial amendments, and the proposed informed consent are reviewed and approved by the institutional review board, independent ethics committee, or research ethics board at each trial site or country, in accordance with national legislation. Approval from the Medical Research Ethics Committee Leiden-Den Haag-Delft was granted in July 2019, with the first patient enrolled in August 2019. The trial is registered under ClinicalTrials.gov Identifier NCT04657120.

Data are collected and maintained using the Castor electronic data-capture system and analysed at the Leiden University Medical Center (the Netherlands). Serious adverse events (SAEs) must be reported immediately to Leiden University Medical Center, as well as to the relevant regulatory authority in the country where the SAE occurred. This reporting requirement remains in effect until the study is terminated. As the trial has been classified as low-risk with no significant concerns regarding patient safety, no Data Safety Monitoring Board is involved in overseeing patient safety throughout the trial.

### Clinical trial sites

In the Netherlands, the enrolling sites include Leiden University Medical Center (Leiden, coordinating centre), Amsterdam UMC (Amsterdam), Bernhoven Hospital (Uden), Diakonessenhuis (Utrecht), Flevo Hospital (Almere), Groene Hart Hospital (Gouda), Maasstad Hospital (Rotterdam), Medical Spectrum Twente (MST) (Enschede), Radboud University Medical Center (Nijmegen), Reinier de Graaf Hospital (Delft), and Tergooi Medical Center (Hilversum). In Switzerland, the active sites are Bern University Hospital (Bern, coordinating site), Lausanne University Hospital (Lausanne) and Geneva University Hospital (Geneva). In France, the active sites are University Hospital of Brest (Brest), University Hospital of Saint-Étienne (Saint-Étienne), Louis Mourier Hospital, Assistance Publique – Hôpitaux de Paris (AP-HP) (Colombes), and European Georges Pompidou Hospital – AP-HP (Paris). In Italy, the active sites are A. Gemelli University Hospital Foundation – IRCCS (Rome) and Milan University Hospital – Ca’ Granda Foundation IRCCS (Milan). In Spain, the active site is Ramón y Cajal University Hospital (Madrid). In Belgium, the active site is University Hospital of Antwerp (UZA) (Antwerp). Of these sites, UZA and MST are no longer active in the study due to logistical constraints.

### Trial statistician

H. Putter, PhD, Leiden, the Netherlands.

### Clinical events adjudication committee

H. ten Cate, MD, PhD, and K. Hamulyak, MD, PhD, Maastricht, the Netherlands.

### Dissemination

The results of this trial will be disseminated through publication in a peer-reviewed research journal, presentations at scientific conferences, and social media and digital platforms. After data collection and data cleaning are completed, de-identified data will be registered in a repository and made available for further research on reasonable request to the corresponding author.

## Data Availability

After data collection and data cleaning are completed, de-identified data will be available for further research upon reasonable request to the corresponding author.

## Acknowledgments

**Belgium** – J. Kwakkel, T. Lapperre, A. Hufkens, W. Spysschaert, I. Verhaegen, Dafne Balemans, C. Stessels (Antwerp); **Canada** – G. Le Gal, K. Vrotniakaite (Ottawa); **France** – E. Rolland, R. Verdet, P. Stephan, P. Bourgeois, Z. Onivogui, P. Dias (Brest); H. Salami, M. Chemouny, H. Helfer, N. Izirouel, S. Hadim, L. Plaisance (Colombes); O. Sanchez (Paris); L. Bertoletti (Saint-Etienne); **Italy** – I. Peters (Rome); G. Ramponi (Milan); **The Netherlands** – B. Appelman, S. Kaashoek (Almere); S. Middeldorp, V.L.B.I. Jansen, A. Strijdhorst (Amsterdam); M.M.C. Hovens (Arnhem); F. Kleijwegt, L.M. Faber, C. Bresser (Beverwijk); Z. Jaderi, A.W. Flikweert, B. van Steen, C. Peeters, M. Maas, M. Daalmans (Breda); D.A. van Rijssel, E.M. van der Weert, C. Haazer, A. de Graaf, B. van Prooijen, E.B. van Druten, E.G. Gortmaker (Delft); K. van Elst, C.A. van Assen, E. Geraedts, M. Nagtegaal, A. ten Heuvel, R. Sluga (Gouda); M. de Vreede, Y. Ende, M.A.M. Stals, I. van der Plas, S. Jagernath (The Hague); C.M.M. de Jong, D. Luijten, F.H.J. Kaptein, S.F.B. van der Horst, J.F. Krommenhoek, R.M.A. Mali, S.N.M. Ter Haar, L.J. op de Hoek, P.L. den Exter (Leiden); S. Moll, M. de Bruijn (Hilversum); E. Klappe, O. Sir (Nijmegen); A. Wei, A. Sanders, B. Raap, H. Peters (Uden); M. Deelen, A. Aikema, H. Zanders, M. Hoogervorst (Utrecht); B. Jones, E.M.G. van Berkesteijn (Rotterdam); **Spain** – W. Briceño; R. Morillo; C. Rodríguez-Calle (Madrid); **Switzerland** – S. Maurer (Bern); N. Koffi Malan Antoine (Geneva); H. Gerhard Donnet, M. Brochu Vez (Lausanne).

