## Supplemental Material for "A study protocol for a randomised controlled trial evaluating the safety and efficiency of the YEARS algorithm versus computed tomography pulmonary angiography only for suspected acute pulmonary embolism in patients with cancer: the Hydra Study"

**Authors’ names (e-mail):**
E.S.L. Martens^1^ (0000-0003-2197-2290;), B. Akerboom^1^, C. Baumgartner^2^, R.E. Brouwer^3^, C. Cavallaro^4^, M. Coppens^5a,b^, G. Costantino^6a,b^, F. Couturaud^7^, A. D’Errico^4^, Y.P.A. Dooren^8^, F. Gianni^6a,b^, R. van der Griend^9^, M.J.J.H. Grootenboers^10^, O. Hugli^11^, T. van der Hulle^12^, Z. Jaderi^10^, D. Jimenez^13a,b,c^, P.W. Kamphuisen^5a,b, 14^, V.R. Lanting^5a,b, 14^, J. Leentjens^15^, M.L. Maas^16^, I. Mahé^17^, O.A. van Meer^18^, T.E. van Mens^1^, M. Out^19^, R. Pola^4^, D. Pulver^2^, J. Raskin^20^, M. Righini^21^, R.A. Sprenger^22^, M.A.M. Stals^1^, R. Talerico^4^, T. Tritschler^2^, M. ten Wolde^23^, F.A. Klok^1^, M.V. Huisman^1^, on behalf of the HYDRA study group^24^.

**Affiliation:**
^1^ Department of Medicine - Thrombosis and Hemostasis, Leiden University Medical Center, Leiden, The Netherlands.

^2^ Department of General Internal Medicine, Inselspital, Bern University Hospital, University of Bern, Bern, Switzerland.

^3^ Department of Internal Medicine, Reinier de Graaf Gasthuis, Delft, The Netherlands.

^4^ Percorso Trombosi, Dipartimento di Scienze dell’Invecchiamento, Ortopediche e Reumatologiche, Fondazione Policlinico Universitario A. Gemelli IRCCS, Università Cattolica del Sacro Cuore, Rome, Italy.

^5a^ Department of Vascular Medicine, Amsterdam UMC, University of Amsterdam, Amsterdam, The Netherlands.

^5b^ Amsterdam Cardiovascular Sciences Research Institute, Pulmonary Hypertension & Thrombosis, Amsterdam, The Netherlands.

^6a^ Emergency Department, Fondazione IRCCS Ca' Granda Ospedale Maggiore Policlinico, Milan, Italy.

^6b^ Department of Clinical and Community Sciences, University of Milan, Millan, Italy.

^7^ INSERM U1304-GETBO, INNOVTE network, Department of Pulmonology, Brest University Hospital and Brest University , Brest, France.

^8^ Department of Pulmonology, Groene Hart Hospital, Gouda, The Netherlands.

^9^ Department of Medicine, Diakonessenhuis Utrecht, Utrecht, The Netherlands.

^10^ Department of Pulmonology, Amphia Hospital, Breda, The Netherlands.

^11^ Emergency Department, Lausanne University Hospital and Lausanne University, Lausanne, Switzerland.

^12^ Department of Oncology, Leiden University Medical Center, Leiden, The Netherlands.

^13a^ Respiratory Department, Ramón y Cajal Hospital (IRYCIS), Madrid, Spain.

^13b^ Department of Medicine, Universidad de Alcalá (IRYCIS), Madrid, Spain.

^13c^ CIBER de Enfermedades respiratorias (CIBERES), Madrid, Spain.

^14^ Department of Medicine, Tergooi Hospital, Hilversum, The Netherlands.

^15^ Department of Medicine, Radboud Institute for Health Sciences, Radboud University Medical Centre, Nijmegen, The Netherlands.

^16^ Department of Medicine, Bernhoven Hospital, Uden, The Netherlands.

^17^ Department of Medicine, Hôpital Louis-Mourier, Colombes, Assistance-Publique-Hôpitaux de Paris, Paris Cité University, INSERM Unité Mixte de Recherche U970, Paris Cardiovascular Research Center, Team "Endotheliopathy and Hemostasis Disorders", Paris, France.

^18^ Emergency Department, Leiden University Medical Center, Leiden, The Netherlands.

^19^ Department of Medicine, Medisch Spectrum Twente, Enschede, The Netherlands.

^20^ Department of Pulmonology and Thoracic Oncology, Antwerp University Hospital, Edegem, Belgium.

^21^ Division of Angiology and Hemostasis, Geneva University Hospital and Faculty of Medicine, Geneva, Switzerland.

^22^ Department of Medicine, Maasstad Hospital, Rotterdam, The Netherlands.

^23^ Department of Medicine, Flevo Hospital, Almere, The Netherlands.

^24^ Clinical Research Unit Internal Medicine, Leiden University Medical Center, Leiden, The Netherlands.

**Acknowledgments:**
**Belgium** – J. Kwakkel, T. Lapperre, A. Hufkens, W. Spysschaert, I. Verhaegen, Dafne Balemans, C. Stessels (Antwerp); **Canada** – G. Le Gal, K. Vrotniakaite (Ottawa); **France** – E. Rolland, R. Verdet, P. Stephan, P. Bourgeois, Z. Onivogui, P. Dias (Brest); H. Salami, M. Chemouny, H. Helfer, N. Izirouel, S. Hadim, L. Plaisance (Colombes); O. Sanchez (Paris); L. Bertoletti (Saint-Etienne); **Italy** – I. Peters (Rome); G. Ramponi (Milan); **The Netherlands** – B. Appelman, S. Kaashoek (Almere); S. Middeldorp, V.L.B.I. Jansen, A. Strijdhorst (Amsterdam); M.M.C. Hovens (Arnhem); F. Kleijwegt, L.M. Faber, C. Bresser (Beverwijk); Z. Jaderi, A.W. Flikweert, B. van Steen, C. Peeters, M. Maas, M. Daalmans (Breda); D.A. van Rijssel, E.M. van der Weert, C. Haazer, A. de Graaf, B. van Prooijen, E.B. van Druten, E.G. Gortmaker (Delft); K. van Elst, C.A. van Assen, E. Geraedts, M. Nagtegaal, A. ten Heuvel, R. Sluga (Gouda); M. de Vreede, Y. Ende, M.A.M. Stals, I. van der Plas, S. Jagernath (The Hague); C.M.M. de Jong, D. Luijten, F.H.J. Kaptein, S.F.B. van der Horst, J.F. Krommenhoek, R.M.A. Mali, S.N.M. Ter Haar, L.J. op de Hoek, P.L. den Exter (Leiden); S. Moll, M. de Bruijn (Hilversum); E. Klappe, O. Sir (Nijmegen); A. Wei, A. Sanders, B. Raap, H. Peters (Uden); M. Deelen, A. Aikema, H. Zanders, M. Hoogervorst (Utrecht); B. Jones, E.M.G. van Berkesteijn (Rotterdam); **Spain** – W. Briceño; R. Morillo; C. Rodríguez-Calle (Madrid); **Switzerland** – S. Maurer (Bern); N. Koffi Malan Antoine (Geneva); H. Gerhard Donnet, M. Brochu Vez (Lausanne).

**Corresponding author:** Emily Sascha Louise Martens, MD; Department of Medicine – Thrombosis and Hemostasis, Leiden University Medical Center, Leiden, the Netherlands; Phone: +31-71-5298096;; ORCiD iD: 0000-0003-2197-2290

### Adjudication of suspected events during follow-up

An independent adjudication committee, consisting of two thrombosis specialists who are not involved in the study and are unaware of the allocated diagnostic management strategy, will evaluate all new and recurrent suspected clinical events during the 90-day (± 7 days) follow-up period. The events include symptomatic and incidental pulmonary embolism (PE), deep vein thrombosis (DVT), or death, and major bleeding (MB).

Documentation required for the adjudication of suspected clinical events is limited to clinical summaries, imaging reports, and other relevant findings, e.g., laboratory or physical exam. Requests for copies of films or images from objective tests will only be made when deemed necessary. Disagreements will be solved by discussion between the two specialists of the adjudication committee. If necessary, a third thrombosis specialist not involved in the study will be asked to resolve any persisting disagreements.

*Venous thromboembolism****:***

PE (acute symptomatic or incidental) is diagnosed as follows:

1. by computed tomography pulmonary angiography (CTPA) showing an intraluminal defect in a subsegmental or greater pulmonary artery; or
2. by ventilation-perfusion scanning indicating a mismatch between ventilation and perfusion with a perfusion defect at a segmental or more proximal level; or
3. with inconclusive results of the aforementioned imaging modalities, but objectively confirmed proximal DVT of the lower extremity (as defined below) in patients with clinical signs and symptoms of PE; or
4. by autopsy showing PE

Lower-extremity DVT (acute symptomatic or incidental) is diagnosed as follows:

1. by compression ultrasound (CUS), including grey-scale or colour-coded Doppler, revealing an area of non-compressibility in a venous segment involving at least the popliteal vein or a more proximal leg vein (i.e., femoral and/or iliac veins); or
2. by ascending contrast-venography showing an intraluminal filling defect in the contrast-filled vein, indicating an obstruction due to thrombus formation; or
3. by magnetic resonance (direct thrombus) imaging showing hypointense thrombotic material in a vein or a high signal when imaged using a T1-weighted magnetic resonance imaging (MRI) sequence; or
4. by autopsy showing bland thrombus in at least the popliteal vein or a more proximal leg vein

Upper-extremity DVT (acute symptomatic or incidental) is diagnosed as follows:

1. by CUS, including grey-scale or color-coded Doppler, revealing an area of non-compressibility in a venous segment involving the subclavian, axillary, brachial, and/or brachiocephalic vein; or
2. by ascending contrast-venography showing an intraluminal filling defect in the contrast-filled vein, indicating an obstruction due to thrombus formation
3. by magnetic resonance (direct thrombus) imaging showing hypointense thrombotic material in a vein or a high signal when imaged using a T1-weighted MRI sequence; or
4. by autopsy showing bland thrombus in the deep veins of the arm

Recurrent VTE is defined as:

1. a new intraluminal filling defect or an extension of an existing defect; or
2. an area of non-compressibility in a venous segment where compression had been normal; or
3. a significant increase (>4mm) in vein diameter of a previous non-compressible segment upon CUS.

*Deaths:*

All deaths during follow-up will be adjudicated as to the likelihood that the death was related to PE using the International Society on Thrombosis and Haemostasis (ISTH) definition for PE-related death and classification of the cause of death^20^:

A. PE-related death (i.e., certainly / highly probable PE-related)

A1. Autopsy-confirmed PE in the absence of another more likely cause of death

A2. Objectively confirmed PE (≥1 of the situations as stated above) in the last 48 hours* before death, in the absence of another more likely cause of death

* Longer time period may apply on a case‐by‐case basis

A3. PE is not objectively confirmed, but is most likely the main cause of death

B. Undetermined cause of death (i.e., possibly PE-related)

B1. Cause of death is undetermined, despite available information

B2. Insufficient clinical information available to determine the cause of death

C. Cause other than PE (i.e., PE-unrelated), including:

1. death related to cancer, defined as death caused by direct effects of a malignancy or death in a subject with progressive cancer who had a gradual decline in general condition or in whom palliative treatment only was decided;

2. cardiovascular death including death from acute myocardial infarction, stroke, congestive heart failure, arrhythmia, cardiac surgery, systemic embolic disease other than PE, or other cardiovascular disease;

3. fatal bleeding defined as a bleeding event leading to death, such as intracranial haemorrhage with subsequent brain herniation;

4. death from infection defined as death following septic shock or death cause by direct effects of an infection such as brain abscess with subsequent brain oedema leading to death;
5. other known cause of death (e.g., renal failure);

6. cause of death unknown but the cause of death was most likely other than PE.

For the main analysis assessing the safety of both diagnostic strategies for ruling out PE, both category A (“PE-related death”) and category B (“undetermined cause of death”) will be considered PE-related.

*Major bleeding*

‘MB’ in non-surgical patients is defined according to the criteria of the ISTH ^18^ as clinically overt bleeding which was:

1) fatal; and/or associated with any of the following:
2) a fall in haemoglobin level of ≥2 g/dL (i.e., ≥1.24 mmol/L); or
3) documented transfusion ≥2 units of packed red blood cells or whole blood; or
4) involvement of a critical area or organ, such as intracranial, spinal, ocular, pericardial, articular, intramuscular with compartment syndrome, retroperitoneal).

‘MB’ in surgical patients is defined as ^17^:

1) fatal bleeding; and/or
2) bleeding that is clinically overt and involves a critical area or organ, such as intracranial, intraspinal, intraocular, retroperitoneal, pericardial, in a non‐operated joint, or intramuscular with compartment syndrome, assessed in consultation with the surgeon; or
3) extra-surgical site bleeding causing a fall in haemoglobin level of ≥2 g/dL (i.e., ≥1.24 mmol/L), or leading to transfusion of two or more units of whole blood or red cells, with temporal association within 24–48 hour to the bleeding; or
4) surgical site bleeding that requires a second intervention – open, arthroscopic, endovascular – or a hemarthrosis of sufficient size as to interfere with rehabilitation by delaying mobilization or delayed wound healing, resulting in prolonged hospitalization or a deep wound infection; or
5) surgical site bleeding that is unexpected and prolonged and/or sufficiently large to cause haemodynamic instability, as assessed by the surgeon. There should be an associate fall in haemoglobin level of ≥2 g/dL (i.e., ≥1.24 mmol/L), or transfusion, indicated by the bleeding, of at least two units of whole blood or red cells, with temporal association within 24 hours to the bleeding.
